# Automated Interictal Epileptiform Discharge Detection From Scalp EEG Using Scalable Time-series Classification Approaches

**DOI:** 10.1101/2022.07.06.22277287

**Authors:** D. Nhu, M. Janmohamed, L. Shakhatreh, O. Gonen, P. Perucca, A. Gilligan, P. Kwan, T. O’Brien, C. W. Tan, L. Kuhlmann

## Abstract

Deep learning for automated interictal epileptiform discharge (IED) detection has been topical with many published papers in recent years. All existing work viewed EEG signals as time-series and developed specific models for IED classification; however, general time-series classification (TSC) methods were not considered. Moreover, none of these methods were evaluated on any public datasets, making direct comparisons challenging. This paper explored two state-of-the-art convolutional-based TSC algorithms, InceptionTime and Minirocket, on IED detection. We fine-tuned and cross-evaluated them on two private and public (Temple University Events - TUEV) datasets and provided ready metrics for benchmarking future work. We observed that the optimal parameters correlated with the clinical duration of an IED and achieved the best AUC, AUPRC and F1 scores of 0.98, 0.80 and 0.77 on the private datasets, respectively. The AUC, AUPRC and F1 on TUEV were 0.99, 0.99 and 0.97, respectively. While algorithms trained on the private sets maintained the performance when tested on the TUEV data, those trained on TUEV could not generalise well to the private data. These results emerge from differences in the class distributions across datasets and indicate a need for public datasets with a better diversity of IED waveforms, background activities and artifacts to facilitate standardisation and benchmarking of algorithms.

## 1. Introduction

Epilepsy diagnosis involves visual analysis of scalp EEG recordings, in which interictal epileptiform discharges (IEDs) are used as biomarkers. IEDs are spike-wave complexes, standing out from background activities. The duration of an epileptic spike varies between 20 and 200 milliseconds.^1^ An IED with repetitive spikes, however, can last several seconds.^1^ Epilepsy is broadly classified as focal or/and generalised. In focal epilepsy, the epileptic activities start in one area and may spread to other areas, while generalised epileptic activities start in both cerebral hemispheres at the same time. A visual example of generalised IED wave-forms is shown in Fig. 1, which can be viewed as a time-series, either multivariate (epoch-wise) or univariate (channel-wise).

**Figure 1:**
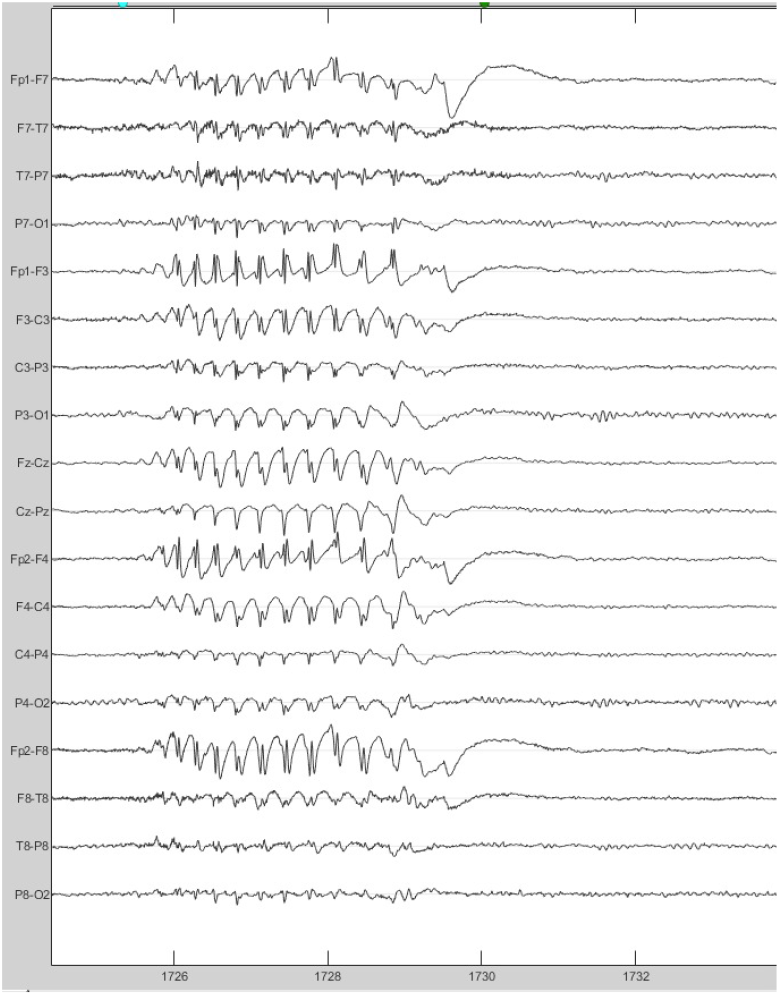
An example of IED from a patient with generalised epilepsy. The IED manifests across all channels.

Manually reviewing EEG recordings to detect IEDs is time-consuming and can take hours to analyse prolonged EEG. Deep learning (DL) has been applied to automate IED detection with promising results in recent years.^2–5^ The 1D convolution neural network (CNN) is the most common architecture.

Traditional machine learning methods have been benchmarked and outperformed by DL approaches;^6^ however, none of the state-of-the-art (SOTA) methods in time-series classification (TSC) were considered. The University of California Riverside (UCR) archive^7^ has been the standard corpus in TSC for benchmarking new models. Numerous studies on this corpus, containing time-series from many disciplines (e.g. finance, health, etc), were published.

Most of the SOTA TSC methods although achieving high accuracy in general TSC tasks, are not suitable for automating IED detection tasks. This is due to the high complexity of the algorithms, with some taking days to weeks to train on a dataset with only 1,000 time series,^8^ while a typical IED dataset contains thousands of time series.^4, 5, 9^

Narrowing down to more scalable algorithms, we deem the InceptionTime^10^ and Minirocket^11^ algorithms to be more suitable for the IED detection task. InceptionTime is an accurate and efficient DL TSC algorithm that can exploit GPU power for large datasets; Minirocket is the fastest TSC algorithm to-date, that is able to complete training and classifying of the whole UCR archive with 109 datasets in less than 10 minutes without sacrificing accuracy.

Existing 1D CNN architectures for IED detection employ a static kernel size in a block of convolution.^2, 9, 12^ However, it was shown in the Inception-Time and Minirocket work with different time-series data that applying different large kernel sizes and concatenating the outputs could achieve better performance than a small kernel size.^10, 11^ This might be the case with IEDs as their duration vary. In terms of public datasets for automated IED detection, there is only one dataset, Temple University Events (TUEV).^13^ This is a subset of a large corpus, Temple University EEG, prepared by Temple University. Unlike the UCR archive, this dataset has not yet earned much attention with only one study having been published to date.^14^

To fill in the gap, we evaluated two SOTA methods from TSC, Minirocket and InceptionTime, on TUEV and provide ready metrics for benchmarking for future work. In addition, the generalisation of these methods was tested on two private datasets acquired at tertiary hospitals in Australia.

## 2. Related Work

### 2.1. Time-series classification

Many studies^15–17^ have been published on general time-series classification since the release of the UCR archive.^7^ HIVE-COTE 2.0^8^ and TS-CHIEF^17^ achieved the best overall performance on the UCR archive. These two methods are ensembles of multiple models from different domains such as the temporal, frequency, shapelet, and intervals. Despite having SOTA results, these models suffer from high computational complexity. With the recent advancements of CNN in different fields (i.e. computer vision, natural language processing), convolutional-based methods, ROCKET, Minirocket, and InceptionTime, have emerged as SOTA approaches to TSC with high scalability. Minirocket^11^ is an optimized version of ROCKET^18^ using a fixed set of kernels. InceptionTime^10^ is a derivative of Inception, a CNN based approach for computer vision, and employs large kernel sizes.

### 2.2. Automated IED detection

Automated IED detection employs the classification of sequential temporal segments of EEG signals into either background activity or containing IEDs. There have been various studies on automated IED detection using general machine learning (ML) methods.^6, 19, 20^ Existing works rely on predefined features encapsulating characteristics of EEG signals, such as time, frequency, and wavelet domains. As the feature selection process is manual, these methods cannot be scaled to large datasets. In general, it has been shown that training on a large dataset would produce better generalisability but require a substantial set of features and, thereby, an expensive feature selection process.^21^

In recent years, DL has received a lot of research interest as an automated feature extraction method for large datasets of EEG recordings for different clinical applications.^22, 23^ Following this success, DL has also been applied to automated IED detection^2–5, 9, 12, 14^ and shown to outperform traditional ML approaches.^6^ The most common architecture is the 1D convolutional neural network (CNN). These were applied on small windows of EEG recordings, aiming to learn temporal features. The application of DL to IED detection is still new and lacks standardization. Despite aiming to extract time-domain features, none of the SOTA in TSC have been tested and used as benchmarks. Furthermore, other clinically relevant metrics, such as F1 or precision, were not reported widely in the literature. The goal of automated IED detection is to reduce the review time, and these metrics would provide an idea of how these models could successfully accomplish this.

### 2.3. Datasets in deep learning for IED detection

The majority of datasets in the literature are private and contain three types of datasets: focal epilepsy, generalized epilepsy, or mixed. Two studies included private datasets on benign rolandic epilepsy with centrotemporal spikes (BECTS), which is a type of focal epilepsy seen only in children.^24, 25^ Generalized epilepsy datasets in existing work are also private and only consist of EEG recordings from patients with idiopathic generalized epilepsy.^2, 3, 26^ TUEV^13^ is the largest public dataset containing both focal and generalised epileptiform activities with different EEG recording settings.^14^ Other studies that experimented with both types of epilepsy included similar private datasets without cross-evaluating on any external sets.^4, 9^ The lack of benchmarking on public datasets makes direct comparisons among studies challenging. In addition, it is not clear in the literature if it is better to build general or specific models for these epilepsy types.

## 3. Methods

Below we outline the datasets considered in this study, preprocessing and data transformation steps, the SOTA TSC machine learning methods employed (InceptionTime and Minirocket), and summarise the different classifier configurations and within- and across-dataset evaluation approaches. Finally we specify the computational implementation details.

### 3.1. Datasets

#### 3.1.1. Temple University Events

Temple University Hospital (TUH) corpus^13^ is a public corpus containing multiple EEG datasets, collected from TUH and managed by Temple University, and released in 2016. The corpus has been updated over time. The TUEV is a subset of this corpus, containing labels of different IEDs, artifacts, and background activities. There are six types of labels: (1) spike and sharp wave (SPSW), (2) generalised periodic epileptiform discharges (GPED), (3) periodic lateralised epileptiform discharges (PLED), (4) eye movement (EYEM), (5) artifact (ARTF) and (6) background (BCKG). One or a set of channels was given labels, and training is then done only using involved channels. This will capture focal abnormalities in maximally involved channels while ignoring channels with artefacts or channels with no abnormalities during the same time window. The duration of all annotations is 1s. TUEV uses the Temporal Central Parasagittal (TCP) montage (Fig. 2). Refer to the original paper of the TUH corpus^13^ for the detailed definitions of these events.

**Figure 2:**
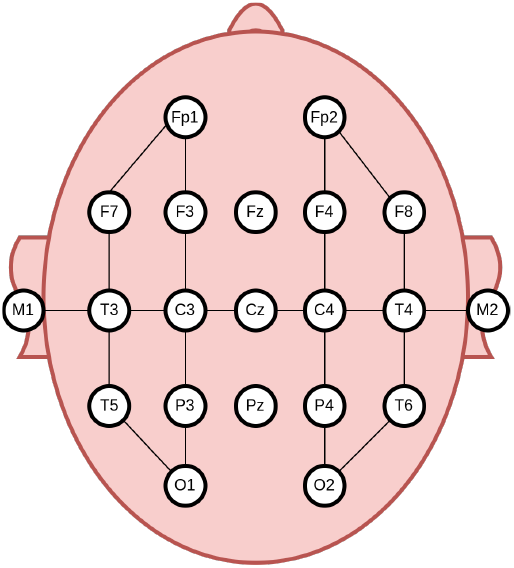
Electrode linkages in the Temporal Central Parasagittal (TCP) Montage. The combined signal from a pair of electrodes was calculated by subtracting one from the other.

TUEV has two sets, train and evaluation sets. These were designed such that data from a patient only appear in one of the 2 sets. We used TUEV version 1.0.1 at the time of this work. Details of the TUEV are summarised in Table 1.

**Table 1:** Number of annotations in TUEV train & evaluation sets

| Annotations | Train set | Evaluation set |
| --- | --- | --- |
| SPSW | 645 | 567 |
| GPED | 11,229 | 4,672 |
| PLED | 5,965 | 1,998 |
| ARTF | 10,194 | 2,204 |
| EYEM | 1,069 | 328 |
| BCKG | 53,126 | 19,602 |
| **Total IEDs** | **17,839** | **7,237** |
| **Total normal** | **65,109** | **22,134** |

#### 3.1.2. Private datasets

Routine EEG recordings from patients with idiopathic generalised epilepsy (IGE) were collected at the Alfred Hospital (n=120) and Royal Melbourne Hospital (RMH; n=137) in Melbourne, Australia. In addition, normal control recordings were obtained from these sites (n=116 and 388, respectively). Three board-certified neurologists (MJ, LS and OG) annotated the datasets. This study was conducted with approval from the Alfred Health Ethics Committee (Project No: 745/19).

Contrary to the channel-wise and 6 class annotation of the TUEV dataset, in the private datasets data epochs were annotated at the epoch level with 2 classes (containing IED or not). We observed that there were fragment IEDs in our datasets which manifest across both hemispheres in some but not all channels in an epoch. The start and end of the spikes for annotation was as follows:

- **Start**: Annotate 100-200 milliseconds before onset of the first visible spike or rhythm change (mark before the first fragment as some discharges start with focal onset in a few channels).
- **End**: End of the last slow-wave and return of predischarge background.

In total, there were 1,155 IEDs in the Alfred Hospital dataset and 1,608 IEDs in the RMH dataset.

### 3.2. Preprocessing

We applied the same preprocessing steps to all datasets. Signals from each electrode were split into 1s windows with no overlap. We standardized the number of electrodes in our dataset by excluding the auricular electrodes M1 and M2, as they were not present in some of the EEG recordings in the Alfred Hospital and RMH datasets. This left us with 19 electrodes for each EEG recording. As the TUEV dataset was labelled with the TCP montage, we applied the same montage to all datasets. All EEG recordings were resampled to 256 Hz using polyphase filtering. We applied bandpass filters of 0.5-49 Hz to remove high-frequency muscle artifacts, slow-frequency artifacts, and power-line noises which do not contribute any information to the diagnosis of epilepsy.^12^ This has been found effective in the literature.^2, 12^ How-ever, the bandpass filtering was not applied in multiclass classification (defined below) as artifacts including high frequency signal content were one of the classes. Finally, the inputs to all models had 256 timesteps and one or 20 channels, for channel-wise or epoch-wise classification (defined below), respectively.

### 3.3. Data transformation

We explored and compared two data transformation methods, z-score normalisation and log transformation. These would help tackle the variance among samples. The data transformation was only applied with InceptionTime. Minirocket does not require any data normalisation as shown in the original paper.

#### 3.3.1. Z-score normalisation

Inputs were z-score normalised with each dataset’s global mean and standard deviation. When training on private sets, the mean and standard deviation were averaged across the two datasets. We also randomly added Gaussian noise to the normalised input with zero mean. The standard deviations were randomly sampled from {1, 2, 3, 4, 5}.

#### 3.3.2. Log transformation

Apart from z-score normalisation, we also explored log transformation. This is independent of the data’s statistics. As the voltage values from our EEG recordings are small/close to 0, we first scaled them up by multiplying with 10^3^. Applying the TCP montage resulted in negative values. Therefore, we squared the inputs and added a small value of 1e-7 for numerical stabilisation. We observed that the distribution of outputs from the log transformation resembled a normal distribution.

In addition, instead of using Gaussian noise for data augmentation in this case, we implemented the mixup method.^27^ Suppose we have two raw inputs *x*_*i*_, *x*_*j*_ and their one-hot encoding labels *y*_*i*_, *y*_*j*_, mixup creates virtual training examples by interpolating these as follows,

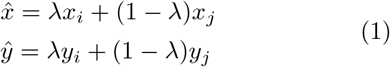

where *λ* ∼ *Beta*(*α, α*) and *α* ∈ (0, ∞). A benefit of this method is it does not rely on the statistics of the dataset. It also penalizes the targets of noisy samples so that the model would not lean towards these too much. In our experiments, we set *α* to 0.2.

### 3.4. SOTA TSC methods

#### 3.4.1. InceptionTime

Inception was first introduced in^28^ and has been through a few iterations of architecture change.^29^ The basic module of an Inception network consists of a concatenation of outputs from different 1D convolutional kernels. InceptionTime^10^ was inspired by this and achieved SOTA results on the UCR archive. The InceptionTime module starts with a bottleneck connection reducing the input features, followed by convolutional operations with different kernel sizes. Differences between InceptionTime and the original Inception are the kernel sizes are longer (64 compared to 3×3 and 5×5), and no pooling layers are used to reduce the input dimensions (Fig. 3). In addition, the residual connection is applied to every two convolutional blocks. We also applied 1D spatial dropout^30^ after every convolutional block to regularize the network. The 1D spatial dropout randomly drops a set of timesteps. In addition, we followed the average ensemble in the original paper. Inception-Time was trained five times with different random seeds, and the outputs of these models were averaged to produce the final output.

**Figure 3:**
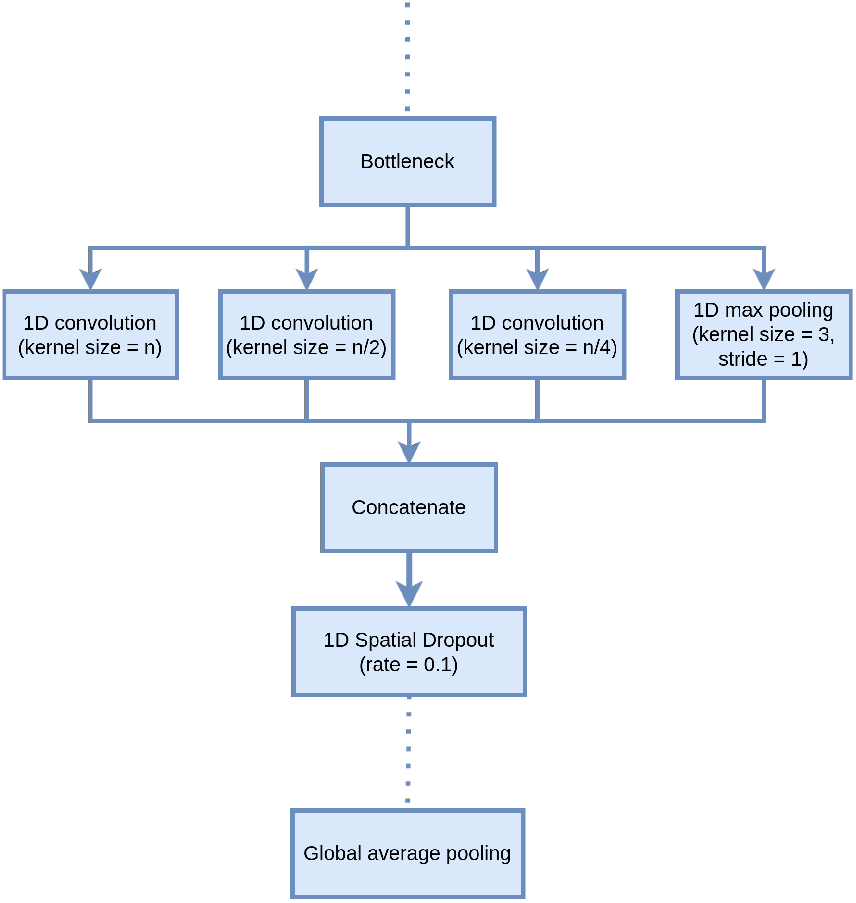
A CNN block of InceptionTime where n is the maximum kernel size. The bottleneck layer is a 1D convolutional layer with kernel size of 1. Each block is followed by a 1D spatial dropout layer in our modification. A global average pooling layer is used to reduce the dimensions before the final classification layer.

#### 3.4.2. Minirocket

Rocket^18^ is a scalable algorithm that achieved SOTA accuracy on the UCR archive. It first transforms the time series using 10k random convolution kernels and computes the proportion of positive values (PPV) statistics and maximum value of the feature maps, creating 20k features. The transformed inputs are then used to train a linear classifier. Minirocket^11^ is an optimised and faster version of Rocket. It uses a fixed set of 10k kernels and only computes the PPV statistics, creating 10k features in total. The convolution in both Rocket and Minirocket uses dilation, indicating the spread of the kernel, to achieve diversity and better accuracy. By default, Minirocket uses the ridge classifier for datasets with less than 10k series and the logistic regression classifier for larger datasets.

### 3.5. Within- and across-dataset evaluation setup

The SOTA TSC methods were evaluated for the TUEV and private datasets separately (i.e. within), as well as across datasets.

#### 3.5.1. Within-TUEV-dataset evaluation

As each label in the TUEV dataset was 1s long, we focused on classifying these 1s epochs/windows. We split the TUEV train set into two sets, train and validation such that no patient existed in both sets. The TUEV evaluation set was used as a test set. The exact numbers of each label are described in Table 2.

**Table 2:** Details of TUEV dataset used for training and evaluation in our experiments

|  | Train | Validation | Test |
| --- | --- | --- | --- |
| # IED channel epochs<sup>†</sup> | 16,776 | 1,063 | 7,237 |
| # Normal channel epochs<sup>†</sup> | 50,580 | 14,529 | 22,134 |
| # IED epochs<sup>††</sup> | 6,319 | 231 | 7,237 |
| # Normal epochs<sup>††</sup> | 7,239 | 1,473 | 22,134 |
| # EEG | 291 | 61 | 159 |
| # Patients | 231 | 57 | 80 |

<sup>†</sup> Epochs of single channel data
<sup>††</sup> Epochs of multichannel data

#### 3.5.2. Within-private-dataset evaluation

In terms of our private datasets, we split each dataset into three sets, train, validation, and test. Each set contained a different set of EEG recordings. We extracted 1s windows with 50% overlap from these recordings. To avoid including any IEDs that the neurologists might have missed, we only used the signal from normal EEG as normal activities. Table 3 contains the details of the data in our experiments.

**Table 3:** The number of epochs of multichannel data and EEG recordings of private datasets

| Alfred Hospital dataset |  |  |  |
| --- | --- | --- | --- |
|  | Train | Validation | Test |
| # IED epochs<sup>†</sup> | 1,024 | 229 | 335 |
| # Normal epochs<sup>†</sup> | 201,629 | 67,640 | 69,424 |
| # IED labels | 827 | 157 | 171 |
| # Epileptic EEG | 58 | 13 | 25 |
| # Normal EEG | 77 | 20 | 24 |
| # Patients | 133 | 33 | 48 |
| RMH dataset |  |  |  |
|  | Train | Validation | Test |
| # IED epochs<sup>†</sup> | 1,371 | 256 | 607 |
| # Normal epochs<sup>†</sup> | 429,726 | 99,166 | 143,007 |
| # IED labels | 1,067 | 165 | 376 |
| # Epileptic EEG | 61 | 16 | 33 |
| # Normal EEG | 255 | 63 | 79 |
| # Patients | 300 | 76 | 111 |

<sup>†</sup> Epochs of multichannel data

#### 3.5.3. Across-dataset evaluation

To test the generalisability of the models trained using the training set of each dataset (i.e. TUEV or the private data), we cross-evaluated the models on the test set from the dataset not used for training (e.g. Train on TUEV train set and test on Private dataset test sets). As these datasets had different types of EEG recordings and epileptiform discharges, this would give us a better understanding of the generalisability of the models and how to better prepare IED datasets for future studies aimed at finding generalisable models.

### 3.6. Classifier Configurations

The goal is to classify windows from EEG recordings into either binary classes, IED or normal, or multiclass as in the TUEV dataset. This work tested SOTA methods in TSC on the automated IED detection task, both channel-wise and epoch-wise. Channel-wise means the classification is performed on channels individually. This can only be done if the events in each channel are labelled by the clinicians. Epoch-wise means the models would be trained either on all channels from a given time epoch or separately on each channel and a voting mechanism is employed to obtain the final classification for the epoch. This requires clinician labelling of events at the epoch level. Moreover, we performed two classification tasks, multiclass and binary classification, which are illustrated in Fig. 4. These approaches were defined as follows:

**Figure 4:**
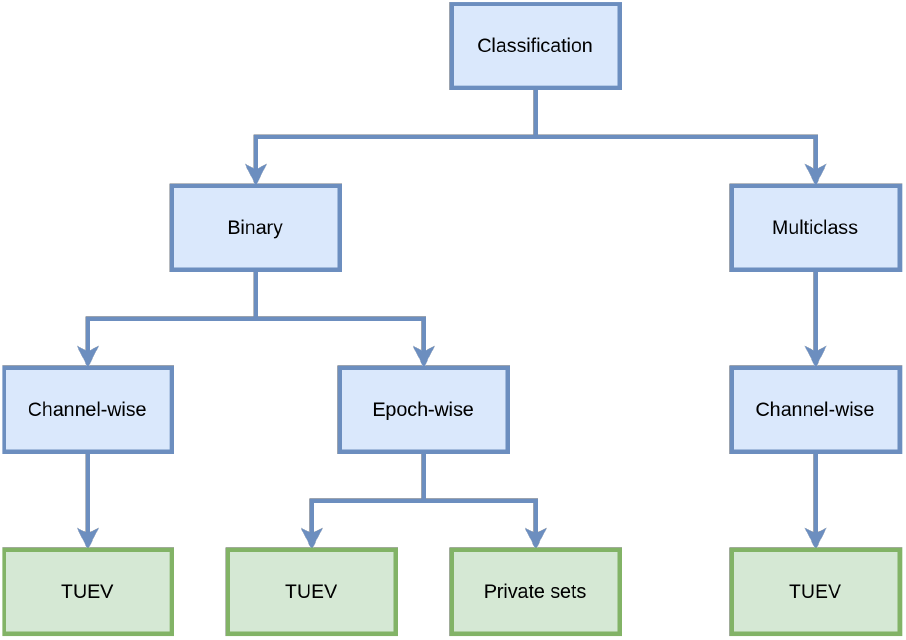
Classification tasks in our experiments. These were performed channel-wise and/or epochwise depending on what labels were available for a given dataset.

#### 3.6.1. Multiclass classification

TUEV dataset had six classes as described above. The multiclass classification was done channel-wise with this set, given that an epoch could contain multiple class labels across the different channels.

#### 3.6.2. Binary classification

For channel-wise binary classification for the TUEV data we grouped SPSW, GPED, and PLED into the IED class and EYEM, ARTF, and BCKG into the non-IED class. For epoch-wise binary classification with the TUEV data, if an epoch contained a channel labelled as SPSW, GPED or PLED, the epoch would be classified as spike, otherwise, it would be classified as normal. To avoid including any channels without IEDs in fragment samples, we only performed epochwise binary classification on the private datasets because events in individual channels were not labelled separately by clinicians and only IED and non-IED class labels were provided.

We explored three strategies for binary classification as seen in literature:^9, 14, 26^

- **Multichannel:** All channels in an epoch were used in training. The inputs were multivariate time-series.
- **Maximum:** We trained the models on binary labels of single channels. The maximum probability output across all channels was considered as the final output of the epoch and used in epoch-wise classifications.
- **Aggregated:** We used the same models from the channel-wise multiclass classification task and aggregated them for epoch-wise classification.

### 3.7. Hyperparameter search

Given the complex hyperparameter space of InceptionTime, we limited the grid search for the optimal hyperparameters to using only the Alfred Hospital private training and validation sets. We set the number of filters in the bottleneck layer to the same value as within the following convolutional layers. We also evaluated whether to use spatial dropout and increase the number of filters every two epochs. The model with the best set of hyperparameters was then used for other experiments, including channel classification. The search space is described in Table 4.The Alfred Hospital dataset was involved during the search and normalised with the z-score normalisation. We employed the average ensemble in which each combination of parameters was trained 5 times with different random seeds and the average of these was used as the final output. Training these models until convergence is costly. To speed up the grid search, we trained each model for 10 epochs and used the AUPRC on the validation set for comparison. We did not perform the hyperparameter search for Minirocket. Instead, we used the default number of kernels of 10,000 in Minirocket and set the maximum dilation to the value of the optimal maximum kernel size of InceptionTime.

**Table 4:** Search values for InceptionTime’s hyperparameters. Optimal values are highlighted in bold.

| Hyperparameters | Search values |
| --- | --- |
| Maximum kernel size | **32**, 40*, 64 |
| Number of filters | 32*, **64** |
| Number of convolutional blocks | 4, 6*, **8** |
| Spatial dropout rate | 0, **0.1**, 0.2 |
| Increase factor for the number of filters | **1**, 1.5, 2 |

\* denotes original value in the InceptionTime paper

The TUEV dataset was imbalanced with a high number of BCKG samples and a low number of SPSW (*<*1%). Our private datasets had the same imbalance problem, with the ratio of IED windows to normal windows of approximately 1:1000. This might make the model biased towards the normal class. The imbalanced dataset is a common issue in deep learning and there have been many solutions.^4, 5, 31, 32^ In our experiments, we employed a balanced mini-batch strategy in which for every batch, we sampled the same number of IED windows and normal windows. This helped create a balanced dataset. All models were trained with a Stochastic Gradient Descent optimizer with weight decay of 1e-5.^33^ We also used the cyclical learning rate^34^ with steps of 8 times the number of batches in one training epoch. The initial and maximum learning rates were 1e-4 and 1e-2.

When working with all datasets, we used the optimal hyperparameters selected from the grid search to train the model for more epochs, maximum of 300, and stopped the training process if the validation set’s AUPRC did not improve after 15 epochs.

### 3.8. Performance Metrics

All models were trained and evaluated on epoched data segments. We used sensitivity to measure the proportion of detected windowed segments containing IEDs. Sensitivity is defined as follows,

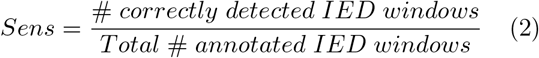

The neurologists will have to verify all suggested samples by the algorithm in practice. To measure the percentage of detected IED windows that are true IEDs, we used precision or positive predictive value which is defined as follows,

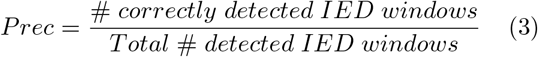

Precision is often traded off against sensitivity. A good algorithm should maintain high precision and sensitivity. We used the F1 score and area under precision-recall curve (AUPRC) obtained for different classification thresholds to measure this. Note recall is the same measure as sensitivity.

F1 score measures the balance between precision and sensitivity and is defined as follows,

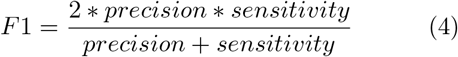

Apart from these, we also computed the area under curve (AUC) of true-positive rate (TPR) and false-positive rate (FPR) at different thresholds. This would give us an idea of the diagnostic ability of the models and is the most commonly used metric in the literature. Noting that the datasets are highly imbalanced, the models might be biased towards the background activities and produce low FPR, resulting in high AUC. As such, the other clinically relevant metrics, F1 and AUPRC, should also be calculated.

### 3.9. Implementation

We extended the original code of InceptionTime, which can be found at https://github.com/hfawaz/InceptionTime, to suit our experiments. We used two implementations of Minirocket from sktime:^35^ univariate and multivariate. All extensions of InceptionTime architecture and logistic regression of Minirocket features were implemented with Tensorflow 2.^36^

## 4. Results

### 4.1. Results of hyperparameter search

The best set of hyperparameters for InceptionTime from the hyperparameter search process are highlighted in bold in Table 4. The maximum kernel size of 32 meant the kernel sizes the model operated on were 8, 16, and 32. As all input windows were 1s long, these are equivalent to 30 to 125 ms and align with the duration of epileptiform waves, which last for 20 to 200 milliseconds.^1^ Following this, we set the maximum dilation of Minirocket to 32 in other experiments. As the optimal hyperparameters correlated with the duration of epileptiform waves, we used the same parameters for training models on TUEV. This would help us understand the differences among datasets in later analysis.

### 4.2. Performance when training with private sets

Considering the case of epoch-wise binary classification we first trained all models on the mixture of the private RMH and Alfred Hospital training sets. We tested the RMH and Alfred Hospital test sets separately to understand the performance difference across the two hospitals. The within-dataset results of InceptionTime and Minirocket are provided in Table 5. The F1, sensitivity and precision scores were calculated at the probability threshold where the validation’s F1 score was maximum. InceptionTime with z-score normalisation had the best average (across the two hospital datasets) AUPRC score of 0.80 and the best average F1 score of 0.77. This was followed by Minirocket with an average AUPRC of 0.79 and an average F1 score of 0.74.

**Table 5:** Alfred/RMH within-dataset epoch-wise binary classification results, and across-dataset results for the TUEV test set.

| Method | Metrics | Alfred | RMH | Avg of private sets | TUEV (only SPSW) |
| --- | --- | --- | --- | --- | --- |
| Inception Time - Z-score | AUC | 0.98 | 0.99 | 0.98 | 0.99 |
|  | AUPRC | 0.72 | 0.87 | **0.80** | 0.76 |
|  | Sens | 0.60 | 0.79 | 0.69 | 0.65 |
|  | Prec | 0.90 | 0.86 | 0.88 | 0.88 |
|  | F1 | 0.72 | 0.82 | 0.77 | 0.75 |
| Inception Time - Log | AUC | 0.97 | 0.96 | 0.96 | 0.90 |
|  | AUPRC | 0.66 | 0.70 | 0.68 | 0.64 |
|  | Sens | 0.49 | 0.75 | 0.62 | 0.54 |
|  | Prec | 0.88 | 0.90 | 0.89 | 0.93 |
|  | F1 | 0.63 | 0.82 | 0.72 | 0.68 |
| Mini rocket | AUC | 0.99 | 0.99 | 0.99 | 0.99 |
|  | AUPRC | 0.74 | 0.83 | 0.79 | 0.78 |
|  | Sens | 0.62 | 0.74 | 0.68 | 0.58 |
|  | Prec | 0.81 | 0.84 | 0.82 | 0.98 |
|  | F1 | 0.70 | 0.78 | 0.74 | 0.73 |

A bootstrap pair-wise statistical comparison with the Welch’s t-test^37^ of F1 scores from these models was also employed. For each pair-wise model comparison, we ran bootstrap 1,000 times and then used the Bonferroni correction for multiple comparisons^38^ to reject the null hypothesis that the means of F1 scores from a given pair of methods were the same. Any null hypothesis with 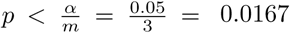 was be rejected, where *m* = 3 is the number of comparisons. We found that all comparisons of the average F1 scores of the models in Table 5 were statistically significant.

We also evaluated across-dataset performance by testing how well these models trained on Alfred Hospital and RMH training sets detected SPSW samples from the TUEV evaluation set. All GPED and PLED samples were removed because these discharge types were not present in the private datasets obtained from IGE patients. As indicated in Table 5, all models generalized well to TUEV test set. The InceptionTime model with z-score normalisation outperformed others with the highest F1 score of 0.75, followed by Minirocket with an F1 score of 0.73.

### 4.3. Performance when training with the TUEV data

#### 4.3.1. Channel-wise Multiclass classification

The within-dataset results of channel-wise multiclass classification on the TUEV evaluation set were summarized in Table 6. We measured the precision, sensitivity and F1 scores of each label. We calculated the macro average (unweighted mean) and weighted average (support-weighted) of all labels to obtain the overall performance. InceptionTime with log transformation had the highest weighted average F1 score of 0.83, followed by Minirocket with a score of 0.77. We observed that all models had trouble detecting SPSW. This could be due to a low number of SPSW samples in the dataset. The Minirocket models had higher F1 scores for SPSW than all InceptionTime models. On the other hand, these models could detect eye movements and artifacts with reasonable precision and sensitivity. This might suggest that these models could be further developed into artifact detection systems.

**Table 6:**
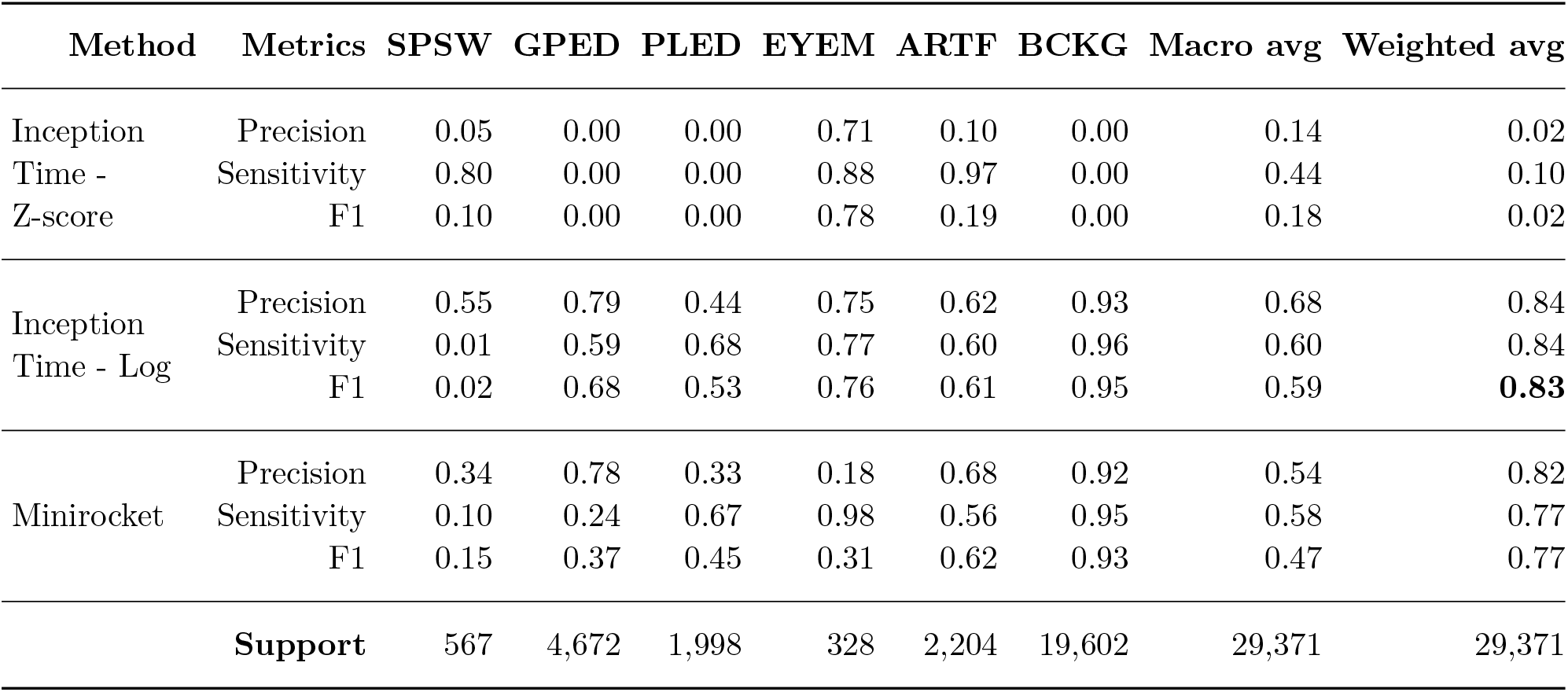
TUEV within-dataset channel-wise multiclass classification results on the test set

In addition, the InceptionTime model with z-score normalisation had low performance. We observed that the model mistook all GPED and PLED samples for SPSW and BCKG samples for ARTF. Note that bandpass filtering was not applied here as we wanted to detect artifacts. This resulted in extremely high variances in the inputs’ scales, accounting for the poor performance. Despite using batch normalisation, the model still failed to distinguish these classes. The proposed log transformation rescaled the inputs, transformed them to a normal distribution, and achieved better results.

#### 4.3.2. Binary classification

This experiment aimed to classify channels and epochs into either normal or containing IEDs. The TUEV within-dataset results of channel-wise binary classification are provided in Table 7. The AUC performance levels of all models are comparable. The InceptionTime model with log transformation had the highest AUPRC and F1 scores of 0.97 and 0.92, respectively. Recall that, regarding epoch classification, we employed three strategies, maximum, multichannel and aggregated for the binary classification. The results of TUEV within-dataset epoch-wise bi-nary classification are reported in Table 8. We did not calculate the AUC and AUPRC for the aggregated strategy as for each sample, we considered the class with the highest probability as the final class and did not apply thresholding. The F1, sensitivity and precision scores of other models were calculated at the threshold where validation’s F1 score was maximum.

**Table 7:**
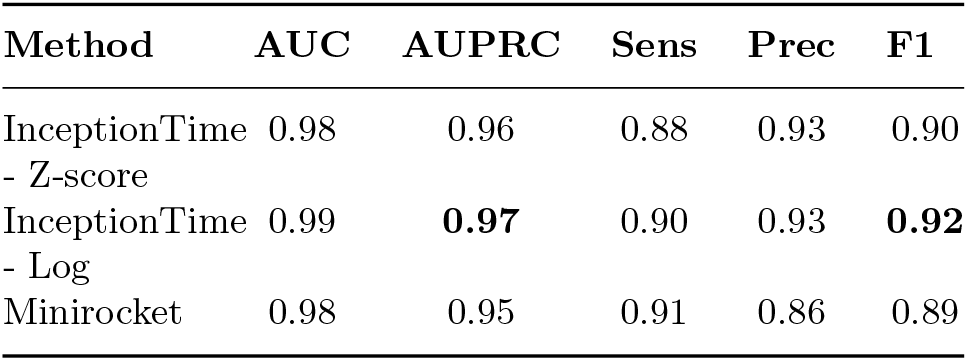
TUEV within-dataset channel-wise binary classification results on the test set

**Table 8:** TUEV within-dataset epoch-wise binary classification results on the test set

| InceptionTime |  |  |  |  |  |
| --- | --- | --- | --- | --- | --- |
| Method | AUC | AUPRC | Sens | Prec | F1 |
| Max - Z-score | 0.99 | **0.99** | 0.92 | 0.96 | 0.94 |
| Multichannel - Z-score | 0.82 | 0.69 | 0.78 | 0.67 | 0.72 |
| Aggregated - Z-score | N/A | N/A | 1.00 | 0.60 | 0.75 |
| Max - Log | 0.99 | **0.99** | 0.98 | 0.95 | **0.97** |
| Multichannel - Log | 0.96 | 0.91 | 0.92 | 0.83 | 0.87 |
| Aggregated - Log | N/A | N/A | 1.00 | 0.82 | 0.90 |
| Minirocket |  |  |  |  |  |
| Method | AUC | AUPRC | Sens | Prec | F1 |
| Max | 0.98 | 0.97 | 0.95 | 0.90 | 0.92 |
| Multichannel | 0.98 | 0.97 | 0.95 | 0.91 | 0.90 |
| Aggregated | N/A | N/A | 0.99 | 0.82 | 0.90 |

All models had comparable AUC scores. InceptionTime with log transformation models had higher F1 scores than respective InceptionTime models with z-score normalisation. We applied the same bootstrap statistical comparisons to confirm this and found the differences were significant. The maximum strategy with the average ensemble outperformed the other strategies. The InceptionTime model with maximum strategy and either z-score normalisation or log transformation shared the best AUPRC score of 0.99. We noticed that the aggregated classification models achieved higher F1 scores in binary classification while struggling to detect individual IED labels. This indicates that all models had trouble distinguishing wave-forms of different IEDs but managed to recognise IEDs in general.

It can also be seen in Table 8, that the InceptionTime with log transformation and maximum strategy had the highest F1 score in terms of epoch-wise binary classification, followed by Minirocket with maximum strategy. We applied the same bootstrap statistical comparison used for the private data to the TUEV data to determine if there were statistically significant differences in the F1 score of given model pairs. Significance occurred if 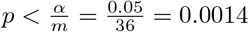 using Bonferroni correction for *m* comparisons. Fig. 5 illustrates the F1 scores and the results of statistical comparisons among all models.

**Figure 5:**
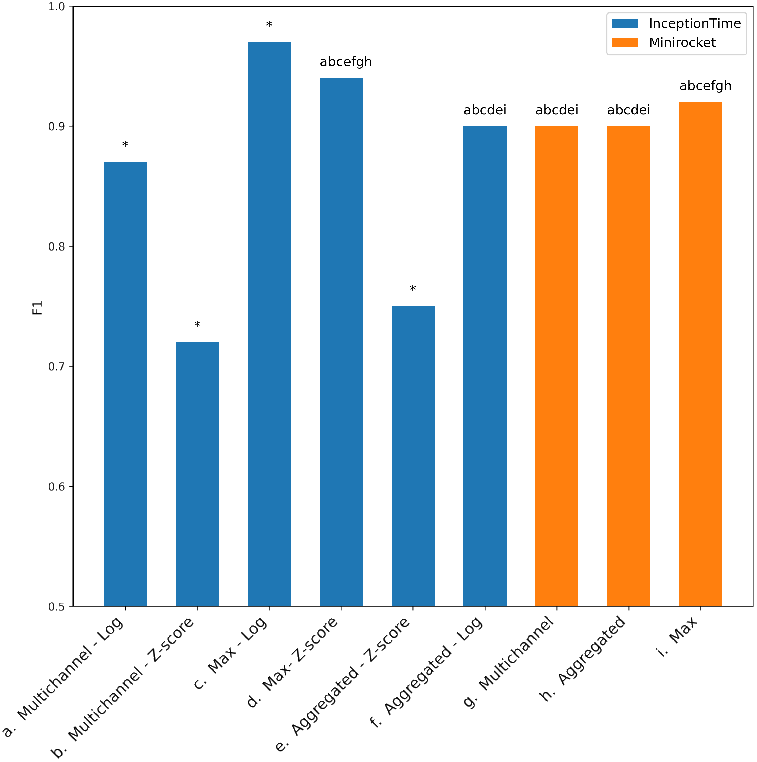
Results of F1 statistical comparisons of TUEV within-dataset epoch-wise Binary-classification. Each method is annotated with a letter between a and i, as seen on the x-axis. Symbols above a given bar indicate if, after correcting for multiple comparisons, there was a significant difference in the F1 score corresponding to the methods associated with the given bar and with the symbol. E.g. a ‘g’ above the bar associated with ‘i. Minirocket - Max’ indicates that ‘i. Minirocket - Max’ and ‘g. Minirocket - Multichannel’ have significantly different F1 scores. An * above a bar indicates that the associated method is significantly different from the rest.

#### 4.3.3. Test on private datasets

We also tested the generalisability of the above TUEV trained models on the test sets from the private Alfred Hospital and RMH datasets. Only binary classification was tested as these sets did not include multiclass labels.

Table 9 summarizes the average metrics across the two private Alfred Hospital and RMH test sets. The AUC scores and sensitivity scores are high, close to the TUEV evaluation set results. However, the precision and F1 scores dropped drastically and were lower than those trained on the private sets. This might be due to the differences between TUEV and private sets. While TUEV contains both focal and generalised IEDs from long-term EEG recordings, the private sets only contain routine EEG recordings from patients with idiopathic generalised epilepsy. There were no PLED and GPED samples in the private sets. We demonstrated in the previous section that these models had trouble detecting SPSW samples from the TUEV dataset.

**Table 9:** Average metrics on private test sets of models trained on TUEV

| InceptionTime |  |  |  |  |  |
| --- | --- | --- | --- | --- | --- |
| Method | AUC | AUPRC | Sens | Prec | F1 |
| Max - Z-score | 0.94 | 0.24 | 0.64 | 0.10 | **0.18** |
| Multichannel - Z-score | 0.91 | 0.03 | 0.96 | 0.02 | 0.03 |
| Aggregated - Z-score | N/A | N/A | 0.98 | 0.01 | 0.03 |
| Max - Log | 0.97 | 0.26 | 0.78 | 0.07 | 0.12 |
| Multichannel - Log | 0.98 | **0.44** | 0.89 | 0.09 | 0.16 |
| Aggregated - Log | N/A | N/A | 0.87 | 0.04 | 0.08 |
| Minirocket |  |  |  |  |  |
| Method | AUC | AUPRC | Sens | Prec | F1 |
| Max | 0.95 | 0.06 | 0.37 | 0.20 | 0.10 |
| Multichannel | 0.98 | 0.37 | 0.91 | 0.08 | 0.14 |
| Aggregated | N/A | N/A | 0.77 | 0.07 | 0.13 |

### 4.4. Exploring false positives

In multiclass classification, the InceptionTime with z-score normalisation classified GPED and PLED samples in the TUEV evaluation set as SPSW. However, the same model performed well for binary classification, where the goal is to distinguish samples with IEDs from any other background and artifact activities. Other models in multiclass classification had the same difficulty in differentiating types of IEDs and performed well in binary classification. On the other hand, while models trained on the private sets generalised well to the TUEV data, we also saw that models trained on the TUEV dataset could not generalise to the two private datasets. Sensitivity scores were high, and precision scores were low.

By analysing the results from the multiclass models, we observed that many background activities in the private datasets were mistaken for either GPED or PLED. This might suggest that there might be differences in the non-IED segments from TUEV and the private datasets, or better artifact removal preprocessing methods are needed. To the understand differences between the TUEV and private datasets, and the lack of generalisation in the train on TUEV case, we explored two common features used in IED detection and artifact removal, spectral entropy^39, 40^ and Hurst exponent.^39, 41, 42^ These features measure the complexity of the signals. Considering the trained on TUEV binary classification results, InceptionTime with z-score normalisation and maximum strategy had the highest cross-evaluation F1 but had the lowest sensitivity. However, we chose the InceptionTime with log transformation and binary multichannel strategy for this experiment as it had the second-best F1 score, and the sensitivity was significantly higher than the other model.

For the two feature types, spectral entropy and Hurst exponent, we compared the feature values corresponding to false positive and false negative epochs detected by this model against the values observed in each of the different classes in the different datasets and plotted box plots in Fig. 6. As multichannel samples were used here, we calculated the feature for each channel of a sample and took the maximum value across all channels as the final value. We implemented the algorithm for Hurst exponent from Qian et al. (2004)^41^ in which the sample was divided into 8 smaller windows of 32 time-steps, similar to the maximum kernel size used throughout our experiments. We implemented the spectral entropy calculation from Pan et al. (2009).^43^

**Figure 6:**
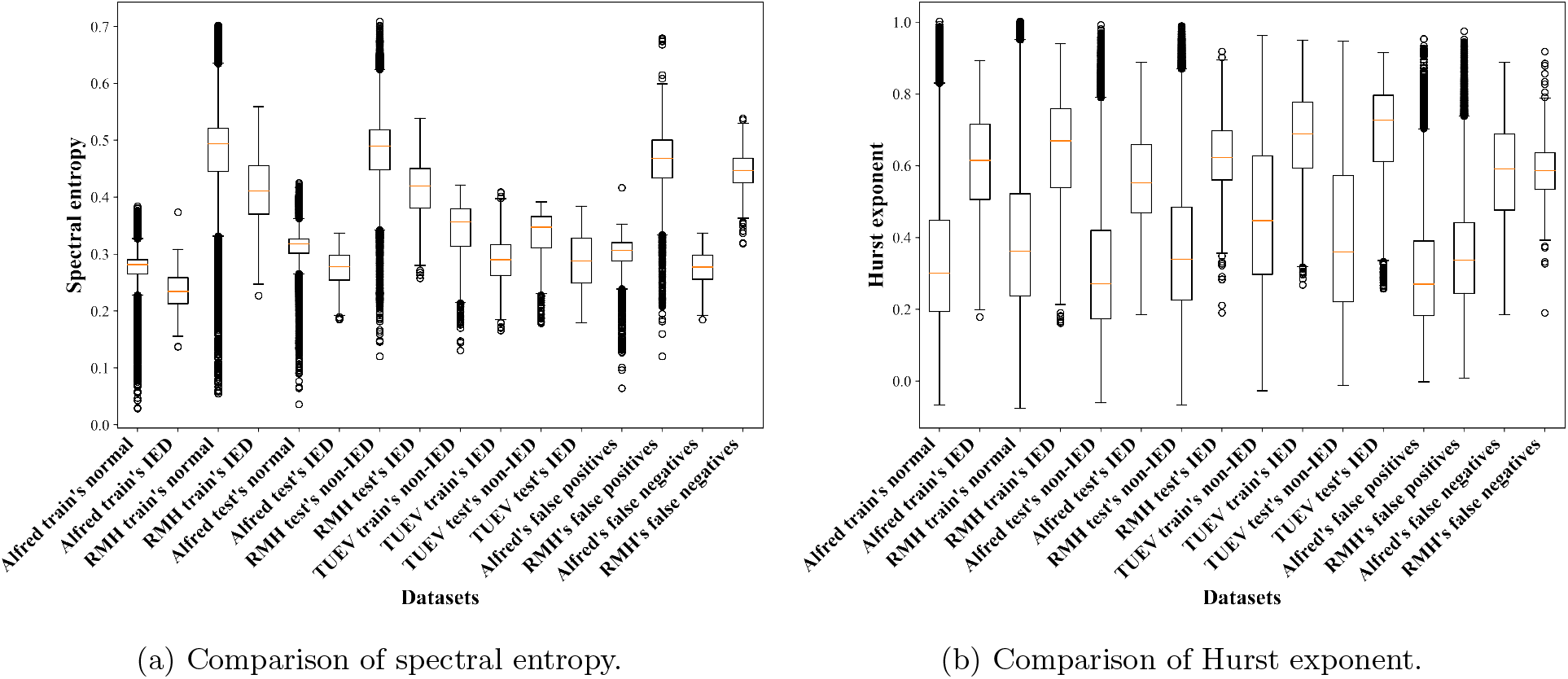
Comparison of features for the TUEV and private datasets, and the false positives and false negatives based on InceptionTime with log transformation and epoch-wise binary multichannel classification.

Even after bandpass filtering, the private datasets were noisier with wider ranges of spectral entropy and Hurst exponent than the TUEV data. This might be partly explained by the much larger number of non-IED examples in the private datasets. The models trained on the private datasets had the benefit of covering more noisy samples, thus, generalized better than those trained on TUEV only. The majority of the false positives from RMH had higher spectral entropy than the IED samples from TUEV. However, spectral entropy for false positives in the Alfred data was close to that observed for TUEV’s IEDs. While there was a large variety of spectral entropy values across datasets (Fig. 6(a)), we observed greater consistency of the Hurst exponent values across datasets (Fig. 6(b)). In particular, it was noticed that a lower bound could be applied to the Hurst exponent values to remove problematic samples and reduce false positives. We tested this by removing any samples from private test sets with Hurst exponent values below different lower bounds between the minimum value and 1st quartile. The IED samples from the TUEV train set were used to calculate the statistics.

The algorithm was as follows: 1. Any samples with Hurst exponent below the lower bound will be classified as normal or without IEDs 2. Let the model classify other samples into either with or without IEDs.

Fig. 7 illustrates the metrics at different lower bounds of the Hurst exponent. We observed that with the trade-off of sensitivity, the precision score improved. This suggests more advanced artifact removal methods might be needed. It would be intuitive to apply this preprocessing step and then train the model on the cleansed data; however, it was out of the scope of this paper where we only wanted to demonstrate the effect of artifact removal and the differences between TUEV and private datasets when using SOTA TSC algorithms.

**Figure 7:**
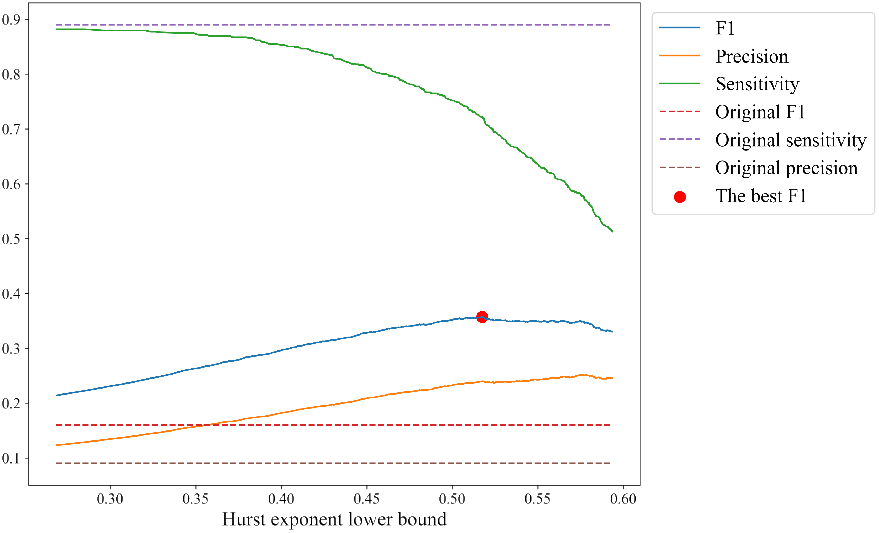
InceptionTime with log transformation and epoch-wise binary multichannel classification metrics after using the Hurst-exponent based algorithm for different Hurst exponent lower bound values.

## 5. Discussion

### 5.1. Overall performance

We evaluated the within- and across-dataset IED detection performance for variants of InceptionTime and Minirocket on our private datasets and the public TUEV dataset. For within-dataset evaluation, the InceptionTime models with log transformation out-performed those with z-score normalisation in most experiments with the TUEV dataset (Tables 6, 7 and 8). By contrast, z-score normalization had better performance on the private datasets (Table 5). This might be due to a wider variation of background activities/artifacts in these datasets that enable the z-score transformation to capture the distribution better. Moreover, Minirocket performed similarly, although not necessarily better, when compared to InceptionTime without the need for normalisation.

When considering across-dataset evaluation work in the literature, the study with data from the most clinical centres (6) is Thomas et al. (2021).^5^ For their proposed model their across-dataset (i.e. across-clinical centre) performance was lower than their within-dataset performance (i.e. within-clinical centre). We experienced the same issue and showed that this was due to differences in the datasets. We demonstrated that the models trained on the private datasets were able to generalise well to the public TUEV dataset (Table 5), while models trained with the TUEV did not generalise well to the private datasets (Table 9). The better generalisation when training on the private sets may result in part from the much larger number of normal/non-IED epochs available in the private sets (Table 3) when compared to the TUEV data (Table 2). Interestingly, we could improve the cross-evaluation performance by using a simple artifact removal method that utilised the thresholding of the Hurst exponent. This might suggest that a better artifact removal method than bandpass filtering is needed. Temple University also released a corpus for artifact detection, TUH EEG Artifact Corpus (TUAR),^44^ which could be used as additional data or to train an artifact detector.

### 5.2. Comparison with existing work

Table 10 compares our work with the literature. Golmohammadi *et al*.^14^ is the only other existing work on the TUEV dataset. This study extracted temporal and spatial context within the EEG data by applying a hybrid method combining Markov models (HMMs) for sequential decoding of EEG events, a deep learning-based post-processing step, and a stacked denoising autoencoder. Only two metrics were reported, sensitivity and specificity, for channelwise binary classification, which were 0.91 and 0.488, respectively. Specificity measures the true negative rate of a classifier. The sensitivity is similar to the best model in our TUEV within-dataset binary classification experiments, InceptionTime with log trans-formation. However, our specificity is significantly higher at 0.98 (see Table 10). In addition, Thomas *et al*.^9^ applied 1D CNN to epoch-wise binary classification on a private dataset with similar EEG recording settings and types of epilepsy to TUEV. The proposed method is similar to our maximum strategy in which the model was trained on single channels, and the maximum probability output across all channels was used as the final output for the epoch. A parameter search was performed over several kernel sizes ranging from 3 to 8, which were smaller than those in our work. The reported AUC was similar to our AUC at 0.99. The AUPRC at 0.84 was lower than our AUPRC at 0.99. Jing *et al*.^4^ employed 1D CNN with increasing kernel size and decreasing number of filters as the network became deeper, and reported an AUC score of 0.98 on a similar dataset to TUEV. Overall, this might indicate that using multiple large kernel sizes could capture epileptic patterns better.

**Table 10:**
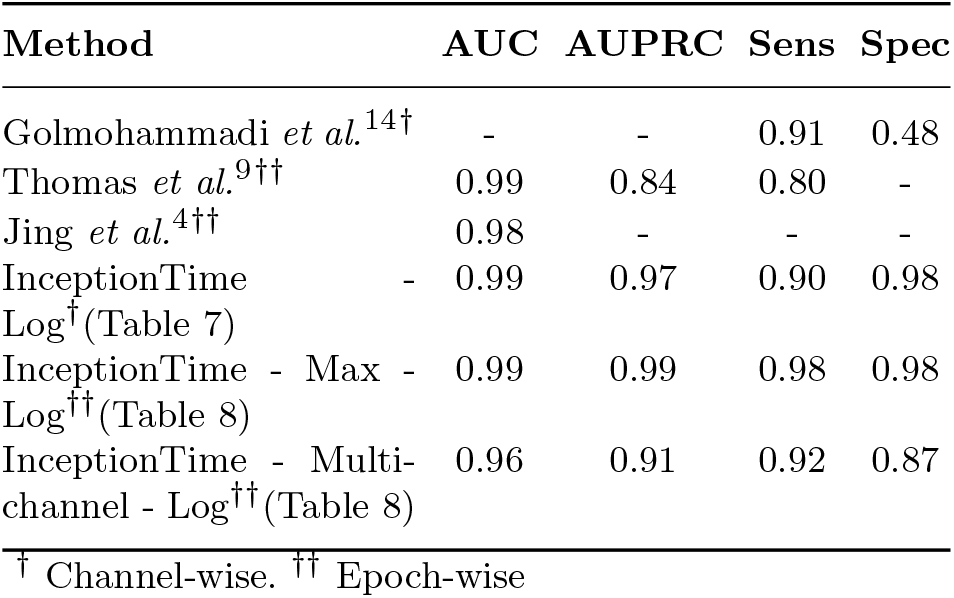
Comparison with existing within-dataset binary classification work on datasets with a mixture of focal and generalized epilepsy patients

In addition to methods that consider both focal and generalised epilepsy patient data as in Table 10, we can also consider the results of considering methods focused purely on IGE patient data, similar to our private data from Alfred Hospital and RMH. Clarke *et al*.^26^ applied 2D CNN and achieved a sensitivity of 0.85. Nhu *et al*.^2^ applied Resnet, a 1D CNN, with different strategies to tackle data imbalance, oversampling, focal loss (FL), and FL with oversampling. The best AUC score achieved was 0.87. Graph convolutional network (GCN) was also experimented with IGE in Nhu *et al*.^3^ which transformed a montage into graph-like data and had an AUC of 0.92 and a sensitivity of 0.60. All methods above used epochwise classification. The comparisons of these results with the performance of the best TSC method for the private data in this paper are given in Table 11.

**Table 11:**
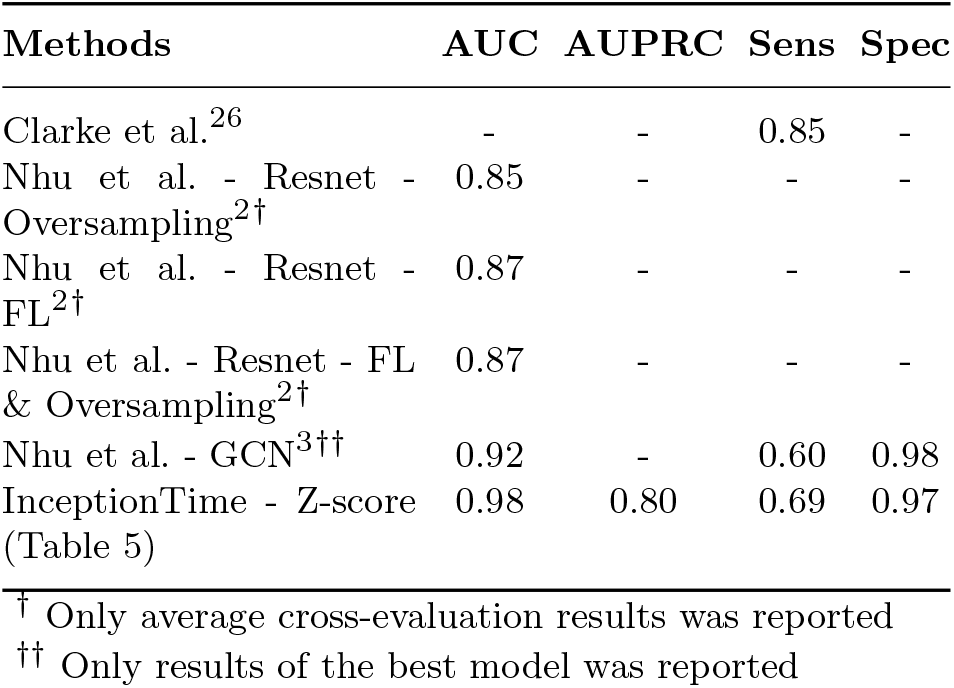
Comparison with existing within-dataset binary epoch-wise classification work on IGE patient only datasets

## 6. Conclusions

This study tested the SOTA TSC methods, InceptionTime and Minirocket, and their variants on the IED detection problem. Ready benchmark scores with relevant clinical metrics on a public dataset, TUEV, were also provided. We also showed that models trained on private data could maintain the performance when tested on the TUEV data. The same was not observed when training on the TUEV data and tested on private data. These results were partly due to the differences in types of IED and normal/artifact activity between TUEV and the private sets. Better generalisation across datasets seems achievable if more normal/artifact data is available for training, as with private datasets. We also showed that our results on the public dataset were comparable in terms of AUC performance and better in terms of AUPRC performance than existing work on DL for automated IED detection. Better AUPRC performance means higher precision, translating into less time wasted by clinicians looking at IED detection that are not actually IEDs. Overall, this work highlights the need for well-structured datasets to improve across dataset generalisation. It also provides SOTA benchmarks for others to compare their algorithms on the public TUEV data.3

## Data Availability

All data produced in the present study are available upon reasonable request to the authors

## Acknowledgment

D.N was supported by the Graduate Research Industry Scholarship (GRIP) at Monash University, Australia. P.K. is supported by a Practitioner Fellowship from the Australian Medical Research Future Fund (MRF1136427). L.K. is supported by NHMRC grants (GNT1183119, GNT1160815).

